# Do improved biomass cookstove interventions improve indoor air quality and blood pressure? A systematic review and meta-analysis

**DOI:** 10.1101/2021.01.04.21249191

**Authors:** Nitya Kumar, Eunice Phillip, Helen Cooper, Megan Davis, Jessica Langevin, Mike Clifford, Debbi Stanistreet

## Abstract

**Background:** Household air pollution (HAP) kills 4 million annually, with access to clean cooking being a challenge for 37% of the world’s population. Whilst there have been advancements in improved biomass cookstove (ICS) technologies, reviews on the impact of these ICS on HAP are now more than three years old.

**Objectives:** This review and meta-analysis examines the most recent evidence on the impact of ICS on HAP and blood pressure (BP).

**Methods:** A literature search was conducted using scientific literature databases and grey literature. Studies were included if they were published between January 2012 and June 2020, reported impact of ICS interventions in non-pregnant adults in low/middle-income countries, and reported post-intervention results along with baseline of traditional cookstoves. Outcomes included 24- or 48-hour averages of kitchen area fine particulate matter (PM_2.5_), carbon monoxide (CO), mean systolic BP (SBP) and mean diastolic BP (DBP). Meta-analyses estimated weighted mean differences between baseline and post-intervention values for all outcome measures.

**Results:** Nine studies were included; eight contributed estimates for HAP and three for BP. Interventions lead to significant reductions in PM_2.5_ (−0.28 mg/m^3^, 95% CI: -0.46, -0.10), CO (−6.59ppm, 95%CI: - 10.73, -2.46) and SBP (−2.82mmHg, 95% CI: -5.53, -0.11); and a non-significant reduction in DBP (−0.80 mmHg, 95%CI: -2.33, 0.73), when compared to baseline of traditional cookstoves. Except for DBP, greatest reductions in all outcomes came from standard combustion ICS with a chimney, compared to ICS without a chimney and advanced combustion ICS. WHO air quality targets were met by post-intervention values for CO but not for PM_2.5._

**Conclusion:** Our review suggests that ICS with a chimney results in the greatest reductions in HAP and BP. Further research on qualitative impact of such ICS on end-users is required to understand feasibility of adoption at scale.

## 1. BACKGROUND

A significant proportion of people residing in low and middle-income countries (LMIC), especially those in the poorest communities, currently do not have access to clean fuel and rely on biomass fuel to meet their daily energy needs for cooking and heating. According to estimates by the IEA (2018), sub-Saharan Africa, specifically, will account for nearly 90% of the world’s population without electricity and gas access and 40% without access to clean cooking by 2030. Traditional cookstoves currently used to burn biomass fuel often lack fuel efficiency and result in incomplete combustion (Bonjour et al. 2013) leading to significantly higher household and ambient air pollution compared to households using clean fuels such as liquified petroleum gas (LPG) and electricity (Yuan et al 2015, Islam et al 2020) Over time, incomplete combustion leads to chronic exposure to fine particulate matter such as that less than 2.5 microns in size (PM_2.5_) and toxic gases such as carbon monoxide (Smith et al 2004, Lea-Langton et al 2019). Both these indicators adversely impact pulmonary and cardiovascular function at all stages of life, and are directly associated with increased incidence of chronic obstructive pulmonary disease, lung cancer, hypertension and stroke, especially in low- and middle-income countries (Bruce et al 2015, Siddharthan et al 2018, Lee et al 2018, Milanzi and Gehring 2019). Exposure to air pollution was estimated to have resulted in 6.5 million deaths in 2015 alone (Global Burden of Disease Study 2015). Of these, 3.8 million result from illness attributable to household air pollution (HAP) caused by the inefficient use of biomass fuels and kerosene during cooking (WHO 2018). To address this disease burden and mortality, it is imperative that a significant reduction in HAP exposure is achieved and to that end, the World Health Organization published guidelines for air quality that set interim targets for PM_2.5_ levels be less than 0.035mg/m3 and the carbon monoxide (CO) levels to be within 6.11ppm (WHO 2014).

Aside from gas and electricity, other effective and cleaner energy alternatives, such as solar and biogas, remain inaccessible and/or unaffordable to the poorest communities. Even with financial assistance, access to, adoption of and sustained use of cleaner fuel alternatives for such communities remains a challenge owing to multiple limiting factors that span across domains of fuel and technology characteristics, knowledge and perceptions, household characteristics, financial tax subsidy aspects, market development and ultimately, regulation and legislation (Puzzolo et al 2016).

In such situations, one of the interim solutions is to enhance existing traditional cookstoves that use biomass fuel (referred to as Improved Cookstoves or ICS) until access to clean energy sources can be achieved in the global poorest communities. So far, improvements in an ICS include adding a chimney feature to the cookstove, refurbishing the combustion chamber to prevent heat loss and finally adding advanced combustion methods that entail enhancing the combustion process by first vaporizing before allowing the combustion to take place (Kshirsagar and Kalamkar 2014).

The reported effectiveness of these ICS from intervention studies over the past decades varies. Some studies report significant improvement in indicators of air quality, such as PM_2.5_ and CO, (Alexander et al 2014, Singh et al 2012, Clark et al 2013) and some reporting no improvements at all (Aung et al 2016, Aung et al 2018). To date, systematic reviews examining air quality improvements of ICS interventions have reported CO reductions from 2.2 to 5.59 ppm and PM_2.5_ reductions ranging from 20 to 490 µg/m^3^(Thomas, et al 2015, Pope et al 2017, Quansah et al 2017). The WHO guidelines (WHO 2014) suggest that even where improved cookstoves have been tested, they are exclusively incapable of reducing emissions to a level close to the interim target. Estimates in these guidelines based on observed concentrations in kitchen field studies, suggest that approximately 60% of traditional stoves had PM_2.5_ concentrations in the range of 500–1800 µg/m^3^ with a mode of 800 µg/m3 and 60% of improved stoves (unvented rocket type stoves) had concentrations in the range of 200–1500 µg/m^3^ with a mode of 500 µg/m^3^. We could consider: Despite the inability of the ICS to achieve WHO targets for PM2.5 and CO in most field studies, most have shown to be better alternatives to the traditional stone fire stoves in terms of emission for communities without access to cleaner source of household energy, especially when used in combination with other interventions such as behavior change communication, among others (Goodwin et al 2015, Stanistreet et la 2014).

Reviews on the impact of ICS on short-term health outcomes from observational studies have reported reductions in COPD (RR=0.74), dry cough (RR=0.72), wheezing (RR=0.41) in adult women (Quansah et al 2017, Thakur et al 2018). Onakomaiya et al (2019) reviewed the impact of ICS on blood pressure from intervention studies carried out with adult women and found SBP reductions ranging from -1.1 to - 5.9mmHg. Conversely, where long-term health outcomes have been measured, the literature indicates that even modest improvements in air quality have the potential to confer long-term health benefits. Shah et al (2015) report in their meta-analysis that for every 1ppm reduction in CO, there was an associated 1.5% reduction in stroke mortality, and every 0.01mg/m^3^ reduction in PM_2.5_ was associated with 1.1% reduction in stroke mortality. Therefore, although none of the reviews report HAP reductions that reached the WHO Air Quality Guidelines target levels, it may be worth promoting an ICS alongside a suite of other interventions that can result in significant improvements in air quality, especially since it may translate to short-term and long-term improvements in health.

To our knowledge only two meta-analyses has explored ICS impact on HAP (Pope et al 2017 and Quansah et al 2017) of which only Quansah et al (2017) provided individual stoves estimates, thereby enabling stove brand recommendation for HAP reducing interventions program. Similarly, we only found one systematic review so far, on the impact of ICS on BP. This study (Onakomaiya et al, 2019) sampled both pregnant and non-pregnant women, making it difficult to draw conclusive inference on the interventions. In addition, these reviews were conducted more than three years ago and have no date cut-off for inclusion of studies. Given these reasons, along with the rapid rate of technology advancements in improved biomass cookstoves (Global Alliance for Clean Cooking 2017, USAID 2017), the present review sought to assess the current best evidence on impact of ICS on HAP and BP, in order to determine which ICS interventions might be most suitable for communities without access to cleaner fuels. This study is part of a larger initiative to identify and recommend an intervention to help HAP and improve proxy indicators of impact of HAP on health, namely, BP, to the poorest of communities in rural Malawi, who are not likely to have access to cleaner fuel alternatives.

### 1.1 Objectives

The main objective of the present review and meta-analysis was to examine the most recent evidence on the impact of ICS interventions on HAP and BP. The secondary objective was to identify which type of ICS was associated with the most promising results and can be recommended to communities that do not have access to cleaner fuels.

## 2. METHODS

### 2.1 Search strategy

A search was conducted on the following databases: Embase, PubMed, Scopus, Web of Science, the BASE, and Global Health Database on OVID. A set of keywords were drafted by DS and EP, with the help of a licensed information specialist and were centered around improved cookstoves, including local terms used to describe them, and household air pollution. The detailed search queries used for each database are provided in the supplementary material (S1). A grey literature search was performed by EP and JL by 1) hand searching references reported in the included studies as well as systematic reviews, which were identified in the initial search; and 2) searching on following websites: Clean Cooking Alliance, Energy for Impact and Berkeley Air Monitoring Group.

### 2.2 Study selection

The search results were exported into Endnote and duplicates were removed. Screening was performed using the application Rayyan and was carried out by 5 reviewers (NK, EP, MD, JL and DS) in a blinded fashion. Each reviewer checked 20% of one other reviewer’s work and any conflicts regarding the exclusion/inclusion of studies were resolved via group discussion and voting. Screening included three stages. Studies were screened based on titles and if required, abstracts, during the first stage. The second stage included screening studies based on abstract and full-text, and the final stage of screening was done at the data extraction stage where studies were excluded if data were not available in the required format for the meta-analysis, either in the published material or from the authors. If data were not available in published documents, authors were contacted. If the data could still not be obtained in the required format, the study was excluded. Since this review came about as a result of preliminary screening of a wider scoping review, it was not prospectively registered; however, the detailed protocol is available upon request.

### 2.3 Inclusion and Exclusion Criteria

#### 2.3.1 Dates of publication

We included studies published between January 2012 and June 2020.

#### 2.3.2 Population

Studies were restricted to those that reported results in adult participants in LMIC in the continents of Asia, Africa and South and Central Americas. Studies reporting impact on blood pressure were excluded if the population included pregnant women given blood pressure fluctuations are common during second trimester and BMI modifies this effect (Rebelo et al 2014), thereby making it difficult to ascertain the impact of ICS on blood pressure in this population.

#### 2.3.3 Interventions

Studies that reported ICS interventions using biomass fuel, were included. We excluded interventions utilizing LPG, solar, biogas and charcoal pellets, which are unlikely to be available and/ or affordable for poor communities, either from fuel procurement perspective or in relation to stove set-up and maintenance, or have been shown not to fully meet cooking needs (Puzzolo 2016)

#### 2.3.4 Comparison

This review sought to compare the post-intervention values of HAP and BP with the baseline values, which corresponded to traditional cookstove use. We, therefore, restricted the studies to those that reported baseline values of outcomes in addition to post-intervention values.

#### 2.3.5 Outcomes

To assess impact on HAP, we considered the outcomes of 24-hour or 48-hour averages of PM_2.5_ and/or 24h/48h CO in the kitchen/cooking area and included studies that examined impact of ICS on these indicators. To assess short-term impact on health, we chose the outcome of blood pressure because it is sensitive to changes in air quality and can help assess impact of short-term interventions better than cardiovascular and pulmonary disease outcomes that have a long induction period and may not reflect short-term impact easily. Studies reporting brachial systolic and diastolic blood pressure measurements were included to assess impact of ICS on BP. We excluded studies where outcomes were less than 24hour averages of PM_2.5_ and CO. Studies that reported modelled or derived values of PM_2.5_ or CO from other proxy variables were excluded.

#### 2.3.6 Study Design

We restricted the review to intervention studies that reported both baseline and post-intervention values of desired outcomes. This criterion was met by randomized controlled trials (RCTs), non-randomized controlled studies and uncontrolled pre-post design studies such as program evaluations. Studies that were lab-based or field-based cooking tests of improved cookstoves, performed only for the duration of the cooking exercise, were excluded.

### 2.4 Data Extraction and Management

Extraction was carried out independently by two reviewers (NK and HC) using the template provided in the supplementary material (S2). Briefly, information on authors, year of publication, location, brand and type of stove, duration of intervention, study design, and outcome data were extracted. Outcome data extracted included means, standard deviations and sample sizes for baseline and post-intervention values in the intervention group. If 95% confidence intervals or median and interquartile ranges were reported, they were converted to means and standard deviations using the methods described by Wan et al (2014). Studies where outcome data were not reported using the above-mentioned summary statistics (e.g., geometric means and percent change in outcomes without actual values for either baseline or post-intervention), and where the data were not provided by the authors on request, were excluded. Wherever data on PM_2.5_ was reported as µg/ m^3^, it was converted to mg/m^3^ prior to analysis.

### 2.5 Risk of Bias

Study quality was assessed independently by NK, EP and HC using the Liverpool Quality Assessment Tool (LQAT) for quantitative studies (Pope et al 2013). Assessment entailed rating the studies based on following five elements: selection procedures (population/sample size, sampling method), assessment of baseline and distribution of intervention, outcome assessment, analysis/ confounding, and applicability/ impact of findings to review. Each element was given a rating of strong, moderate or weak. If none of the five elements had a weak rating, the study was categorized as strong, if a study had one weak rating, it was categorized as moderate, in all other cases, the studies were categorized as weak. An example of assessment of risk of bias is provided in the supplementary material (S3).

### 2.6 Statistical analysis

Data were analyzed using the open-source meta-analysis software from Cochrane Collaboration, Review Manager, version 5.4.1. The effect measure used to assess impact of cookstove intervention on PM_2.5_, CO, systolic blood pressure (SBP) and diastolic blood pressure (DBP) was weighted mean difference between baseline and post-intervention values of the outcome measures. This was done to aid in interpretation of the results in terms of absolute reduction from the intervention as opposed to standardized mean differences. Weighting was done using the inverse variance approach and random effects, as described by DerSimonian and Laird (1986). Heterogeneity between the estimates was assessed using I^2^ using the method described by Higgins and Thompson (2002). Sensitivity analyses were done wherever more than one study or estimate was available for a given outcome measure; these analyses were based on presence of randomization in the study, and duration of intervention and presence of chimney feature in interventions. Statistical significance was assessed using 95% confidence intervals for effect estimates and p values for heterogeneity.

## 3. RESULTS

### 3.1 Overview of included studies

The study search across the databases yielded 1298 results with a final total of 818 studies following removal of duplicates, as detailed in the PRISMA flow chart, shown in Figure 1. Following title and abstract screening, 727 studies were excluded because they were either not related to HAP, did not have an intervention or were not based in low or middle-income countries. From the resulting 91 studies, 41 were excluded on full text screen as they did not include the outcomes of interest or were initial publications from trials, whose updated data was already included from newer studies. Data extraction was performed on 50 studies, out of which, another 41 were excluded when it was found that these studies did not measure baseline values or the reported data was not in a format that was possible to be included in the meta-analysis and, the data in required format was not obtained from the authors upon request.

**Figure 1.**
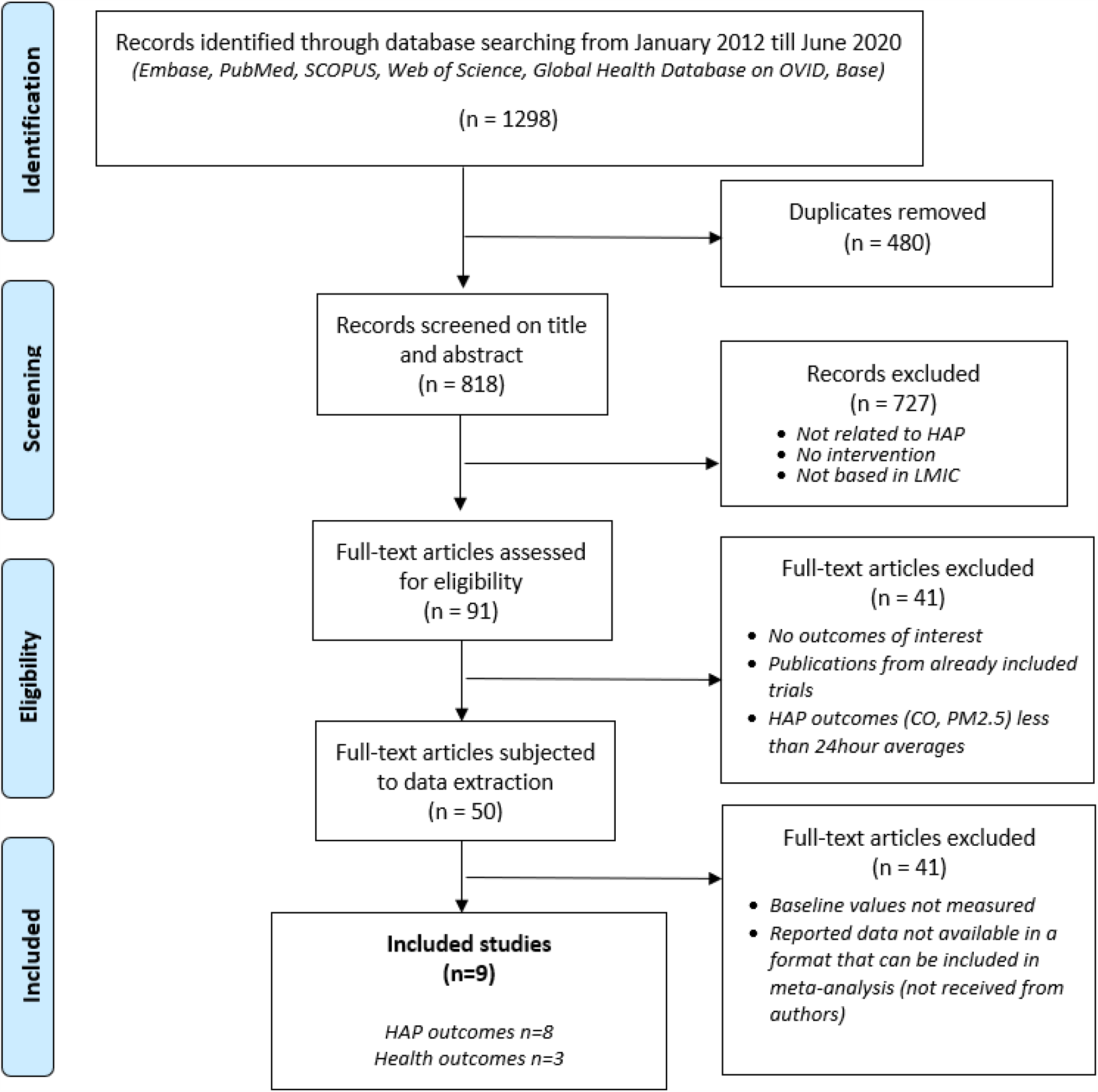
Preferred Reporting Items for Systematic Reviews and Meta Analyses (PRISMA) Indicating Identification and Selection of Studies (Moher et al 2009) Abbreviations: HAP: Household Air Pollution, LMIC: Low- or Middle-Income Countries, CO: Carbon Monoxide, PM2.5: fine Particulate Matter 2.5 microns or less in size

Ultimately, 9 studies were included in the review and analysis, out of which 8 contributed to analysis on HAP outcomes and 4 contributed to analysis on BP. Most of the included studies were from Asia (6), South and central Americas (2), and Africa (1) (Table 1). Only 2 of the 9 studies were RCTs (Aung et al 2016 and Aung et al 2018), 1 was a program evaluation (Clark et al 2019) and the rest were uncontrolled pre-post design studies. The duration of follow-up for all the studies was a minimum 28 days. All the included studies reported estimates for one ICS, except for Sambandam et al (2014) with six different ICS estimates. Except for Sambandam et al (2014), which had 12 participants in their intervention arms, all other studies had at least 20 participants in the intervention arm.

**Table 1.**
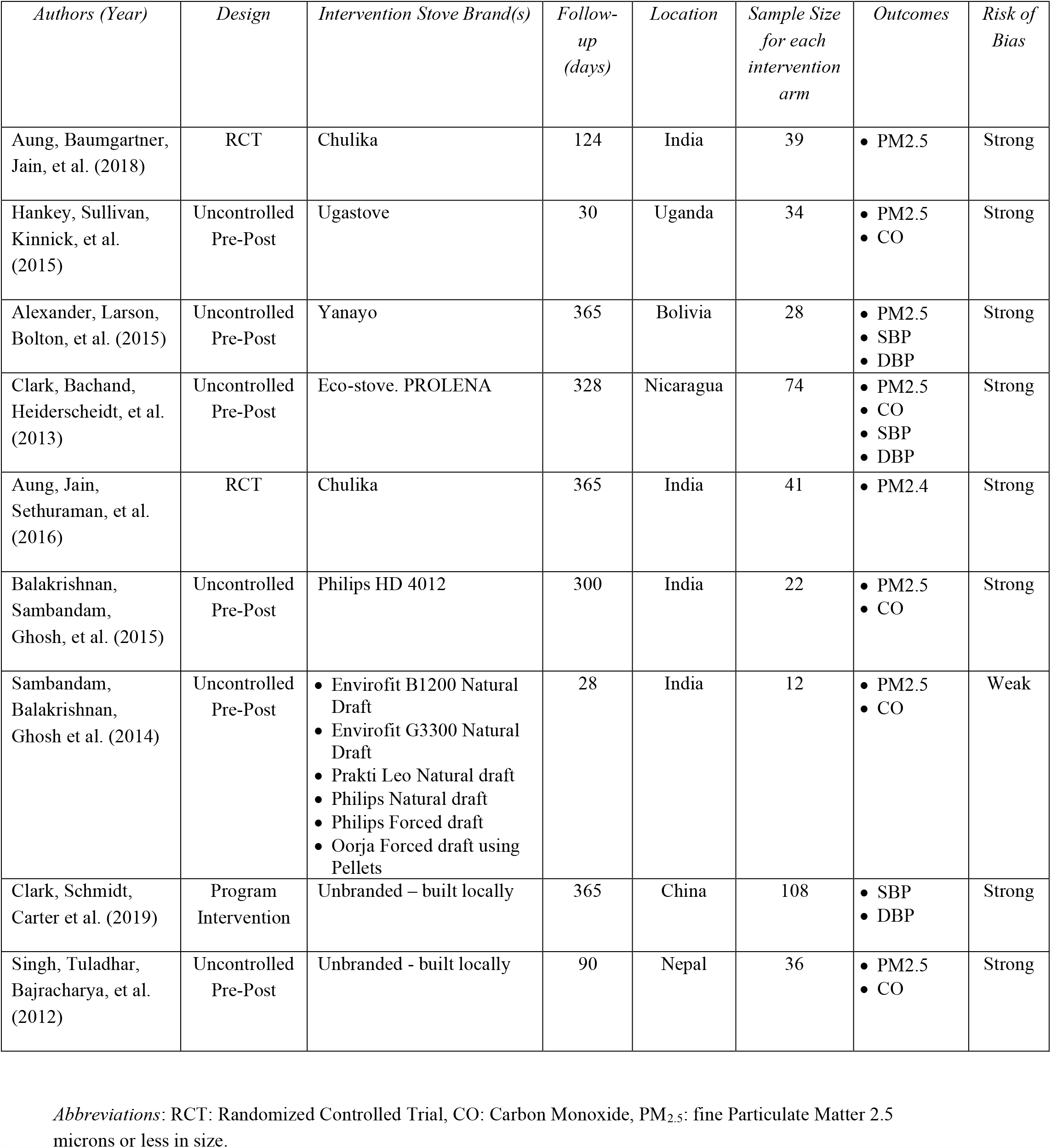
Characteristics of Included Studies.

| *Authors (Year)* | *Design* | *Intervention Stove Brand(s)* | *Follow-up (days)* | *Location* | *Sample Size for each intervention arm* | *Outcomes* | *Risk of Bias* |
| --- | --- | --- | --- | --- | --- | --- | --- |
| Aung, Baumgartner, Jain, et al. (2018) | RCT | Chulika | 124 | India | 39 | • PM2.5 | Strong |
| Hankey, Sullivan, Kinnick, et al. (2015) | Uncontrolled Pre-Post | Ugastove | 30 | Uganda | 34 | • PM2.5<br>• CO | Strong |
| Alexander, Larson, Bolton, et al. (2015) | Uncontrolled Pre-Post | Yanayo | 365 | Bolivia | 28 | • PM2.5<br>• SBP<br>• DBP | Strong |
| Clark, Bachand, Heiderscheidt, et al. (2013) | Uncontrolled Pre-Post | Eco-stove. PROLENA | 328 | Nicaragua | 74 | • PM2.5<br>• CO<br>• SBP<br>• DBP | Strong |
| Aung, Jain, Sethuraman, et al. (2016) | RCT | Chulika | 365 | India | 41 | • PM2.4 | Strong |
| Balakrishnan, Sambandam, Ghosh, et al. (2015) | Uncontrolled Pre-Post | Philips HD 4012 | 300 | India | 22 | • PM2.5<br>• CO | Strong |
| Sambandam, Balakrishnan, Ghosh et al. (2014) | Uncontrolled Pre-Post | • Envirofit B1200 Natural Draft<br>• Envirofit G3300 Natural Draft<br>• Prakti Leo Natural draft<br>• Philips Natural draft<br>• Philips Forced draft<br>• Oorja Forced draft using Pellets | 28 | India | 12 | • PM2.5<br>• CO | Weak |
| Clark, Schmidt, Carter et al. (2019) | Program Intervention | Unbranded – built locally | 365 | China | 108 | • SBP<br>• DBP | Strong |
| Singh, Tuladhar, Bajracharya, et al. (2012) | Uncontrolled Pre-Post | Unbranded - built locally | 90 | Nepal | 36 | • PM2.5<br>• CO | Strong |

*Abbreviations:* RCT: Randomized Controlled Trial, CO: Carbon Monoxide, PM<sub>2.5</sub>: fine Particulate Matter 2.5 microns or less in size.

#### 3.1.1 Stove categorization

The broad objective of this review was to identify and recommend ICS that result in maximum improvements in HAP and BP for communities that do not have access to cleaner fuels. However, most of the included studies focus on ICS that are either locally-made are regional brands available in Asia and South America, with the exception of Philips and Envirofit. Therefore, in order to enable a brand non-specific sub-group analysis and recommendation that is generalizable to communities without access to cleaner fuels globally, ICS were categorized based on their key design characteristics as follows:

1. Standard Combustion Cookstoves with Chimney
2. Standard Combustion Cookstoves without Chimney
3. Advanced Combustion Cookstoves

### 3.2 Impact on HAP

A total of 8 studies (Aung et al 2016, Aung et al 2018, Alexander et al 2015, Balakrishnan et al 2014, Clark et al 2013, Hankey et al 2015, Sambandam et al 2014 and Singh et al 2012) provided estimates for HAP outcomes. Of these, 7 were found to be low risk of bias, and one (Sambandam et al 2014) was found to have high risk of bias because the analysis did not seem to account for kitchen variability and differences at baseline among the intervention arms (Table 1).

#### 3.2.1. Kitchen area PM_2.5_

Overall, the ICS interventions resulted in a significant reduction in kitchen area PM_2.5_ compared to the baseline of traditional stove usage, amounting to –0.28 mg/m3 (−0.46, -0.10), as shown in the forest plot and accompanying table in Figure 2. The estimate was found to have significant statistical heterogeneity at 79% (p<0.001). Sub-group analysis according to stove type, revealed that standard combustion cookstoves showed the maximum mean reduction in post-intervention kitchen area PM_2.5_ values compared with baseline, corresponding to -0.73mg/m^3^ (95% CI: - 1.17, - 0.3). Mean reduction in advanced combustion cookstoves was non-significant at -0.10 mg/m^3^ (95% CI: - 0.34, 0.14, p=0.43). Standard combustion cookstoves without a chimney failed to result in average decline in kitchen area PM_2.5_ values, with the mean change being an increase of 0.05 mg/m^3^ (95% CI: -0.03,0.13, p=0.43) from baseline. Overall, improved cookstoves resulted in a significant reduction in kitchen PM_2.5_ values with most reduction driven by standard combustion cookstoves with chimneys.

**Figure 2.**
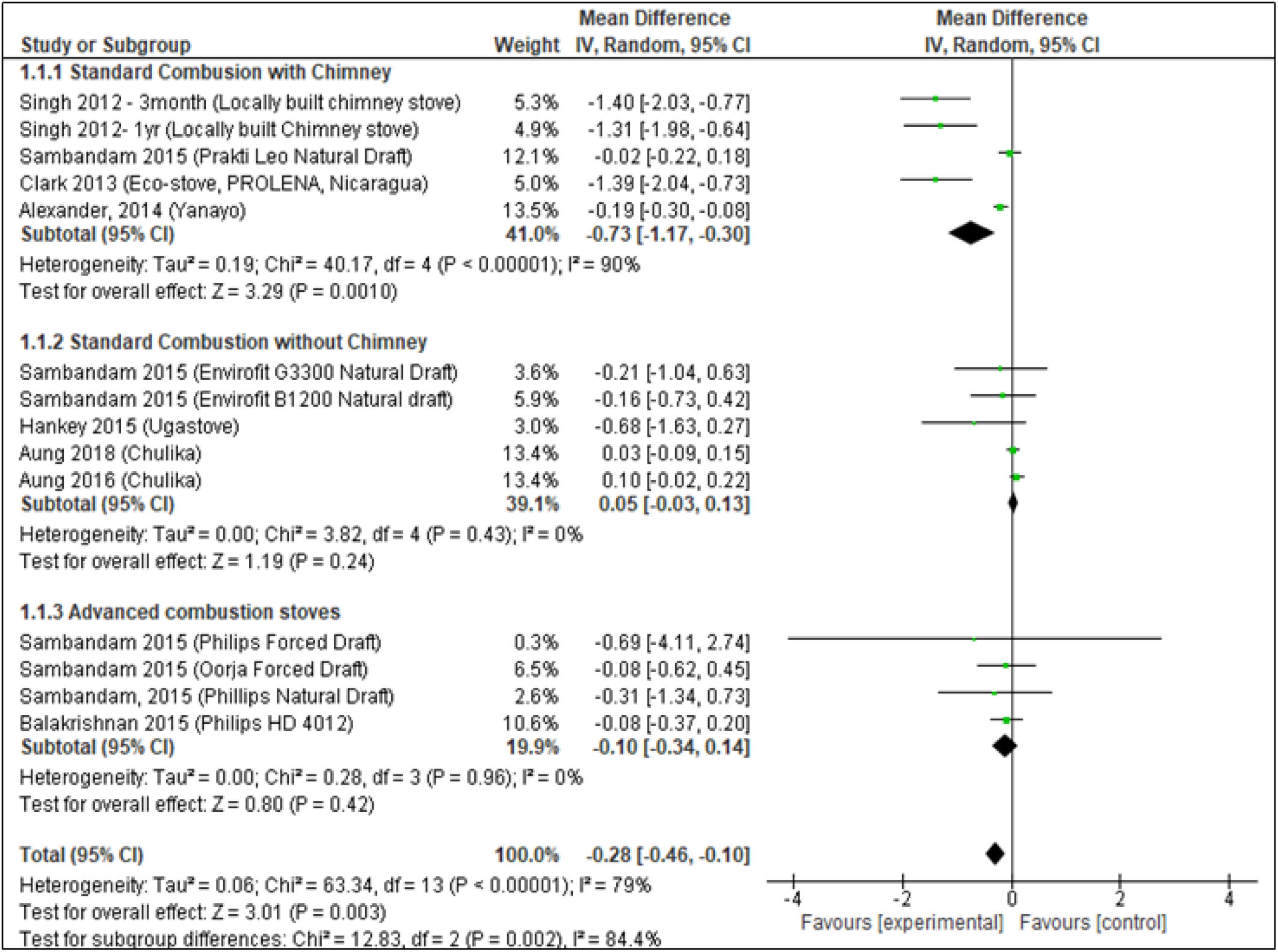
Weighted Mean Differences in Baseline and Post-Intervention Kitchen PM2.5 (mg/m3) Values Across Stove Types.

#### 3.2.2 Kitchen area CO

Comparison of mean CO values before and after intervention with improved cookstoves showed a significant overall reduction of -6.59ppm (95%CI: -10.73, -2.46) as seen in Figure 3. Sub-group analysis according to stove type showed similar trends to those with PM_2.5_. Reductions in CO for standard combustion stoves with chimney were the highest at -14.46 ppm (95% CI: -18.58, -10.34). Standard combustion stoves without a chimney resulted in a mean reduction of -2.01 ppm in CO (95% CI: -7.0, 2.98), and advanced combustion stoves resulted in a similar non-significant reduction of -2.07 ppm (95%CI: -5.02, 0.87, p=0.17). For kitchen CO measurements as well, most of the effect seemed to be attributable to standard combustion stoves with chimney.

**Figure 3.**
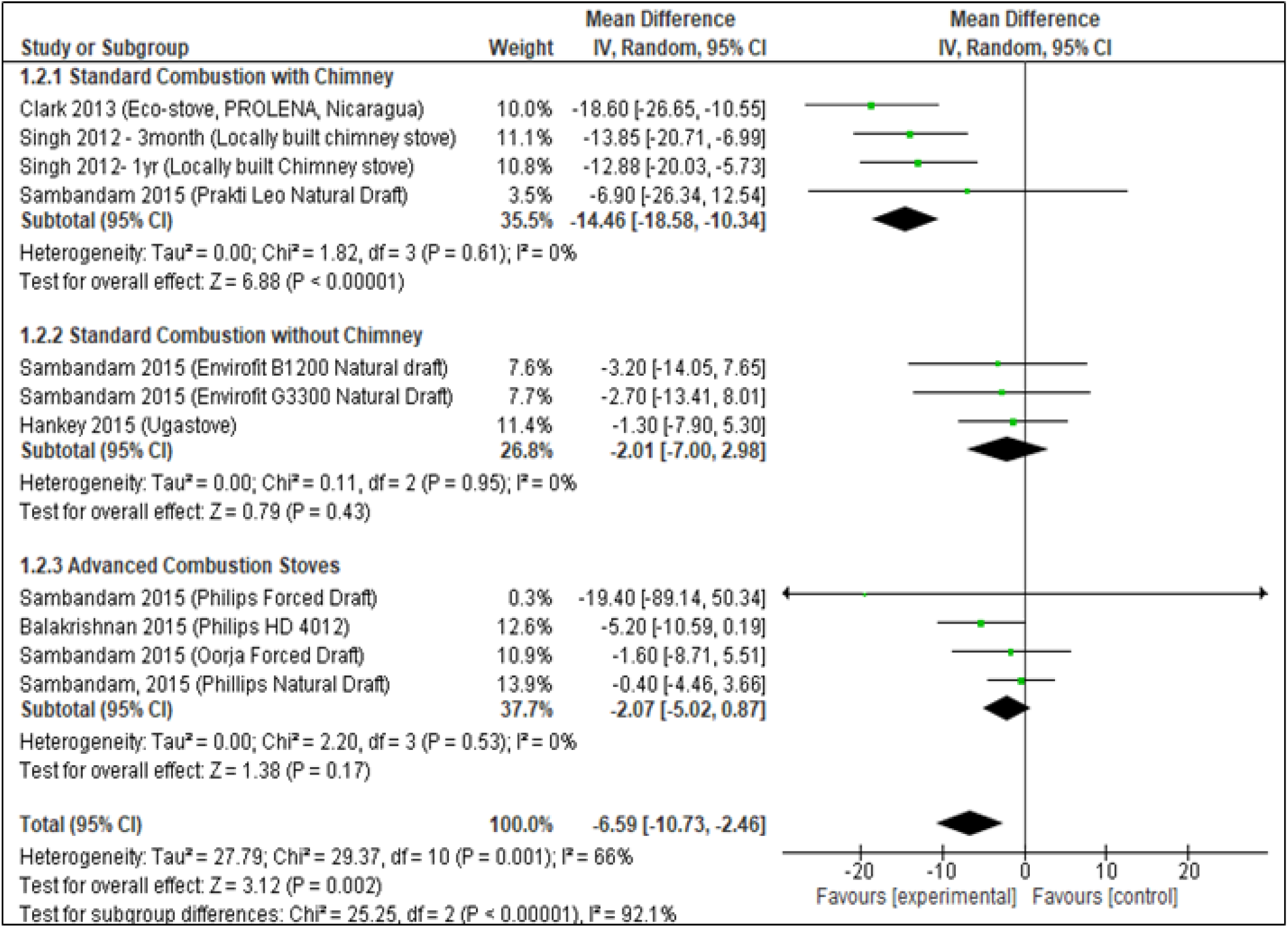
Weighted Mean Difference in Baseline and Post-Intervention Kitchen CO (ppm) Values Across Stove Types.

### 3.3 Sensitivity analyses

Sensitivity analyses were done only for HAP outcomes due to lack of sufficient studies contributing data on blood pressure. These analyses focused on seeing how presence of randomization, duration of follow-up and presence of chimney in stoves affected the estimates for PM_2.5_ and CO, as shown in Table 2.

**Table 2.** Summary of Sensitivity Analyses of Weighted Mean Differences between Baseline and Post-intervention Values of PM2.5 and CO.

| **Sub-group** |  | **PM<sub>2.5</sub> (mg/m<sup>3</sup>)** |  |  | **CO (ppm)** |  |
| --- | --- | --- | --- | --- | --- | --- |
|  | *# of Studies*<br>*(# of estimates)* | *Weighted Mean*<br>*difference (95%*<br>*CI)* | *Heterogeneity -*<br>*I<sup>2</sup> (p value)* | *# of Studies*<br>*(# of estimates)* | *Weighted Mean*<br>*difference (95%*<br>*CI)* | *Heterogeneity -*<br>*I<sup>2</sup> (p value)* |
| **Presence of Chimney** |  |  |  |  |  |  |
| With chimney | 4 (5) | - 0.73 (- 1.17, - 0.3) | 90% (<0.001) | 3 (4) | -14.81 (-19.96, - 9.65) | 0% (0.41) |
| Without chimney | 3 (7) | -0.14 (-0.35, 0.07) | 0% (0.95) | 3 (7) | -1.57 (-3.73, 0.58) | 0% (0.87) |
| **Design** |  |  |  |  |  |  |
| Randomized | 2 (2) | 0.06 (-0.02, 0.15) | 0% (0.44) | 0 (0) | - | - |
| Non-randomized | 6 (12) | -0.44 (-0.69, - 0.19) | 74% (<0.001) | 5 (11) | -5.78 (-9.6, -1.97) | 70% (<0.001) |
| **Duration** |  |  |  |  |  |  |
| 1 month or less | 2 (7) | -0.08 (-0.24, 0.09) | 0% (0.89) | 2 (7) | -1.28 (-4.13, 1.56) | 0% (0.99) |
| Longer than a month | 6 (7) | -0.38 (-0.64, - 0.13) | 90% (<0.001) | 3 (4) | -12.17 (-17.98, - 6.37) | 66% (0.03) |
| **Overall effect** | 8 (14) | - 0.28 (-0.46, - 0.10) | 79% (<0.001) | 5 (11) | -5.78 (-9.6, -1.97) | 70% (<0.001) |

#### 3.3.1. Kitchen area PM_**2.5**_

All three of the above characteristics significantly affected the mean difference in PM_2.5_ It was found that the effect of interventions was most pronounced in ICS with chimney compared to those without a chimney; in non-randomized studies compared to RCTs, and in studies with a follow-up time longer than one month compared to shorter intervention periods. Restricting the analysis to these criteria (chimney present, follow-up>1 month and non-randomized studies), increased the effect estimate to –1.04 mg/m^3^, 95% CI: -1.85, -0.22 (data not shown in tables), which is the highest compared to all other sub-group analyses. However, the confidence intervals were wider and there was no reduction in heterogeneity (I^2^=91%, 0.001). Heterogeneity for all three analyses ranged from 74% to 90% and was not significantly lower than that for the main sub-group analysis based on stove categories (I^2^=79%, Figure 1).

#### 3.3.2. Kitchen area CO

For CO, sensitivity analyses were carried out for presence of chimney and duration of follow-up and not for presence of randomization because none of the randomized studies contributed data on CO (Table 2). Similar to PM_2.5_, the effect estimates for CO reduction were significantly higher for stoves with chimney and in studies with follow-up duration of longer than a month. Restricting the analysis to these characteristics gave a slightly larger mean difference of –14.82 (95% CI: -19.03, -10.6), while the confidence intervals remained mostly similar. As with PM2.5, heterogeneity remained similar in all the CO sensitivity analyses (66%) and was comparable to that observed with the sub-group analysis based on stove categories. Therefore, we choose the stove categorization as the main sub-group analysis and also because it ties to the objective of determining what kind of stove resulted in the most improvement in air quality and blood pressure.

### 3.4 Impact on Blood Pressure

#### 3.4.1 SBP

Systolic blood pressure showed an overall significant reduction of -2.82mmHg between baseline (95%CI: -5.53, -0.11) and intervention with improved cookstoves (Figure 4). Sub-group analysis across stove types, although non-significant, showed similarities with effect on HAP indicators in that the standard combustion stoves with chimney showed the maximum reduction (−3.24, 95%CI: -7.30, 0.83, p=0.12) compared to those without chimney (−2.10, 95%CI: -7.55, 3.35, p=0.45) and advanced combustion stoves (−2.82, 95%CI: -7.69, 2.09, p=0.26).

**Figure 4.**
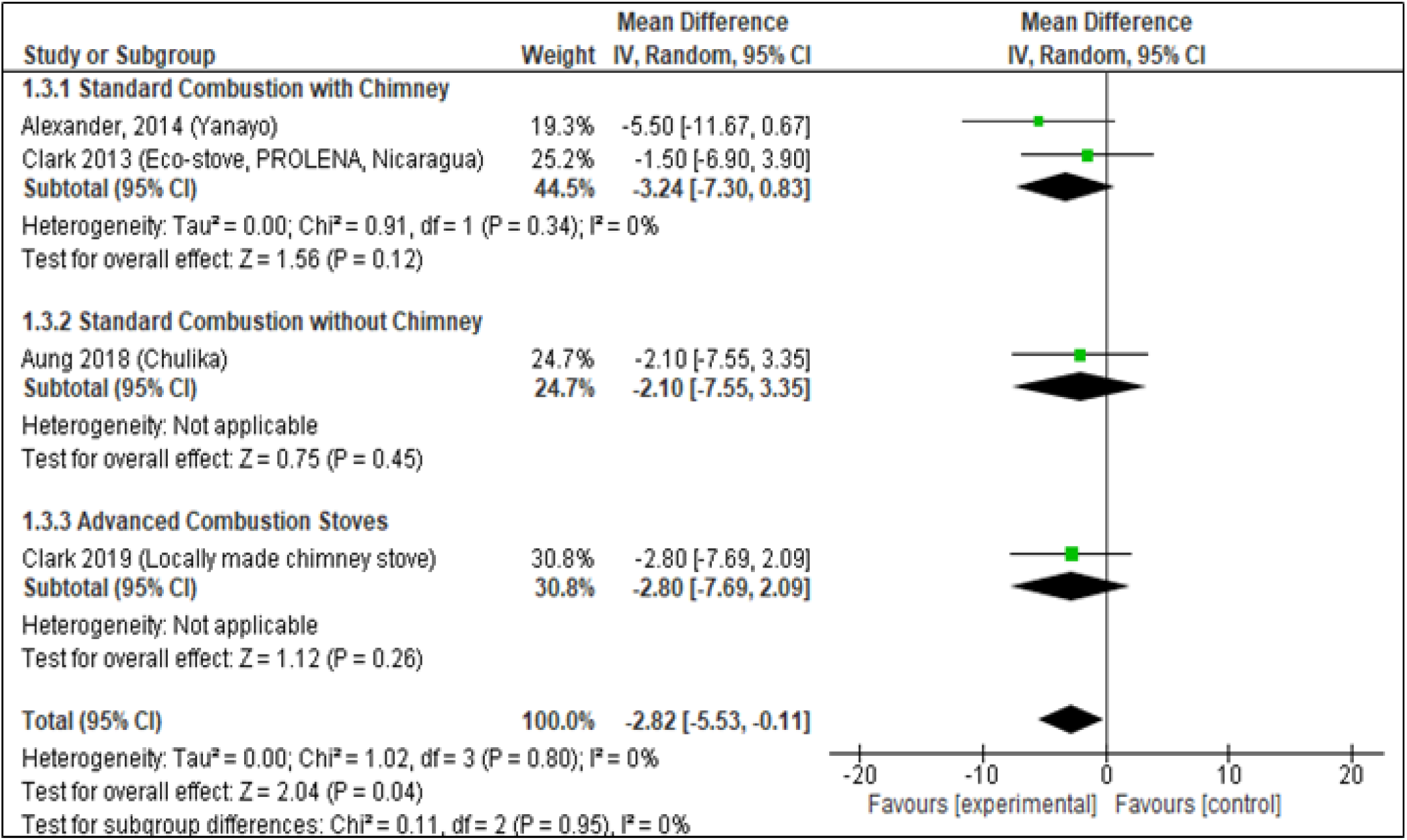
Weighted Mean Difference in Baseline and Post-Intervention SBP (mmHg) Values Across Stove Types.

#### 3.4.2 DBP

Compared to indoor air quality and systolic blood pressure, diastolic blood pressure did not show a significant overall difference between baseline and intervention with improved cookstoves (−0.80mmHg, 95%CI: -2.33, 0.73, p=0.30), as shown in Figure 5. In addition, unlike PM_2.5_, CO and SBP, the standard combustion stoves with chimney showed the least reduction in DBP (−0.44mmHg, 95%CI: -2.80, 1.91, p=0.71) compared to the other types of stoves (standard combustion without chimney = -1.20mmHg, advanced combustion stove =-1.0 mmHg). Ultimately however, the number of studies included in the sub-group analysis is not sufficiently high to draw conclusions based on differences between sub-groups.

**Figure 5.**
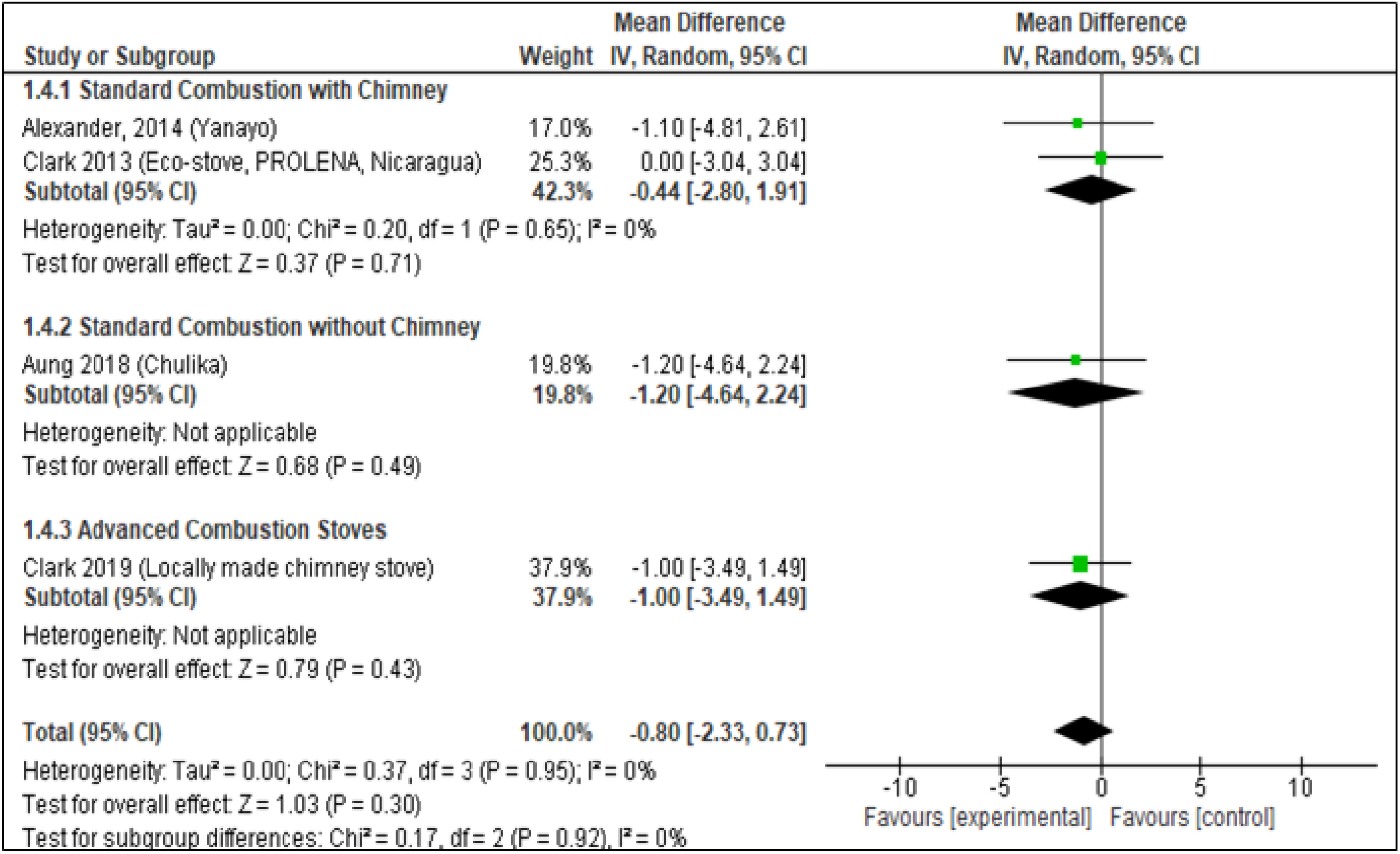
Weighted Mean Difference in Baseline and Post-Intervention DBP (mmHg) Values Across Stove Types.

## 4. Discussion

### 4.1. Kitchen area PM_2.5_ and CO

This review sought to assess latest evidence on the impact of ICS on HAP and health indices, with the main focus being on interventions that would be available to the poorest communities in LMIC. We found that ICS using biomass fuel resulted in an average –0.28mg/m^3^ reduction in PM_2.5_ compared to baseline from traditional cookstoves – equivalent to a 46% reduction in the weighted PM_2.5_ values.

Stoves that had a chimney feature had the highest impact amounting to –0.73 mg/m^3^. With respect to CO, there was an average reduction of –6.59ppm from baseline, which amounts to a 49% reduction from baseline. Stoves with chimney had the highest reduction of 14.46ppm, similar to the findings with PM_2.5_. This finding was also evident in sensitivity analyses where, the greatest reduction in PM_2.5_ was found in interventions with chimney, with a follow-up of more than a month and where studies were non-randomized. If ICS with a chimney do indeed result in most improvements in HAP, then it follows that increasing duration of such interventions would be associated with increased HAP reduction. RCTs included in the review were of shorter duration and hence couldn’t capture this improvement as well as other studies that had longer follow-up.

Although the overall reductions in PM_2.5_ were significant, the post-intervention values for all the studies ranged between 1.16mg/m^3^ to 0.048 mg/m^3^, and none of them met the WHO air quality guideline interim target of 0.035mg/m^3^. The study that reported post-intervention values closest to the guideline (0.048 mg/m^3^) is by Alexander et al (2014) and the intervention was a standard combustion stove with chimney Singh et al (2012) and Clark et al (2013) with baseline PM_2.5_ values of more than 1.5 mg/m^3^, saw the greatest reductions of more than 1 mg/m^3^. All other studies with baseline values of less than 1.5 mg/m^3,^ saw reductions that were less than 1 mg/m^3^. This trend of PM_2.5_ reduction being proportional to the baseline values and the inability of interventions to reduce post-intervention PM_2.5_ values to meet the WHO Guidelines interim target was also evidenced by Thomas et al (2015), Pope et al (2017) and Quansah et al (2017).

Unlike PM_2.5_, the post-intervention values of CO for were found to be within the WHO interim target of 6.11 ppm for 4 ICS interventions (Balakrishnan et al 2015, Sambandam et al 2014). Among these interventions, two (Phillips forced draft and Oorja forced draft cookstove) had low baseline values which themselves were within target (<6.11ppm) and the mean reduction in these groups was modest (−0.40ppm and –1.60ppm). The remaining two stoves that brought the post-intervention values down from above the cut-off to within cut-off, included a standard combustion chimney stove (Prakti Leo Natural Draft cookstove, Sambandam et al 2014) and an advanced combustion cookstove (Philips HD 4012, Balakrishnan 2015). The mean reduction in CO seen in these two ICS (Prakti Leo Natural draft: -6.9ppm -5.2ppm and Philips HD 4012: -5.20ppm were substantially higher, thereby indicating greater impact.

Our findings on standard combustion cookstoves with a chimney producing the greatest reductions in kitchen area PM_2.5_ and CO (–0.73 mg/m^3^ and –6.59ppm respectively) from baseline are similar to the meta-analysis reported by Pope et al (2017) of -0.49 mg/m^3^ and -5.59ppm respectively. The authors also reported considerable heterogeneity (PM_2.5_: 87% and CO: 86%) similar to those in our study (PM_2.5_: 79%, CO: 70%). The only other meta-analysis on impact of ICS on HAP by Quansah et al (2017) found that Plancha stoves resulted in 1.58 standardized mean difference (SMD) of PM_2.5_ in kitchen area compared to Justa stove with 1.00 SMD. Both stoves have a chimney feature. The authors compared these stoves with the rest of the interventions, which included a heterogenous mix of advanced combustion stoves, ethanol, behavior change, and charcoal. Collectively, all of these showed the most PM_2.5_ reduction of 1.68 SMD. This study also found highly significant statistical heterogeneity for kitchen PM_2.5_ (98.2%) and CO (99.5%). The inclusion of a variety of study designs including observational studies, before-after studies without a control group, and RCTs is a common feature in our study as well as Pope et al (2017) and Quansah et al (2017), which could partly explain the heterogeneity.

### 4.2. Blood pressure

The pooled analysis in this study showed a significant reduction of -2.82 mmHg SBP and a non-significant reduction of –0.83mHg in DBP. Restricting the analysis to ICS with chimney revealed higher reductions in SBP of –3.24mmHg and a lower reduction in DBP of –0.44mmHg. These findings are supported by concurrent improvements in air quality reported by two studies in this review. Clark et al (2013) and Alexander et al (2014) reported significant decreases in PM_2.5_ of –1.39 mg/m^3^ and – 0.19mg/m^3^ respectively. In addition, Clark et al (2013) also reported a significant reduction in CO post-intervention amounting to –18.60 ppm. Clark et al (2019) was the only study that did not investigate the impact of the ICS intervention on air-quality. Overall, it appears that the accompanying reductions in PM_2.5_ and CO in these three studies corroborate reduction in SBP. Impact of blood pressure variations during pregnancy can be ruled out here because we restricted the studies on blood pressure to non-pregnant women.

Other factors that could have modified the effect of ICS interventions on blood pressure, include age, BMI, exposure to tobacco smoke, presence of a heart condition or exposure to hypertensive medication. Of the three studies included in the analysis, the maximum SBP reduction was reported by Alexander et al (2015) of –5.50 mmHg. This could be partly explained by the age distribution of the participants in the study, most of whom were older women with a mean age of 51 years. Most of these participants however, were not obese, as indicated by a mean BMI of 23kg/m^2^ and had very low exposure to first- or second-hand tobacco smoke (Alexander et al 2014). The authors did not report presence of hypertension or exposure to hypertensive medications. Clark et al (2013) reported the lowest SBP reduction of –1.5mmHg from baseline. In their sub-group analysis, however, they report an SBP reduction as high as -5.9 mmHg (95% CI: −11.3, −0.4) in women above 40years of age and a reduction of 4.6 mmHg (95% CI: −10.0, 0.8) in obese women. The proportion of participants who had a diagnosed heart condition in this study was 10%. Clark et al (2019) also saw a significant reduction in brachial SBP, which again could be partly explained by the participants’ mean age of 51 years as well as mean a BMI of 24 kg/m^2^, combined with the fact that 60% of participants had exposure to second hand smoke. Similar to Alexander et al 2014, only 14% of the participants were on hypertensive medication, which is unlikely to have affected the outcome significantly. Overall, it appears that older women with higher BMI tend to see most reductions in SBP from ICS interventions due to higher baseline SBP. This is consistent with the evidence synthesized by Onakomaiya et al (2019), the only systematic review until now, that has studied the impact of ICS on blood pressure although their population was also included both pregnant and non-pregnant women.

Stove stacking or the use of multiple types of stoves to meet household cooking and lighting needs has been associated with higher concentration of PM_2.5_ and CO (Yip et al 2016). With the exception of Alexander et al 2014, all other studies included in this review reported presence of stove stacking. This is likely to have attenuated the observed effect of ICS on HAP, as also found by Stanistreet et al (2014) and Rehfuess et al (2014). This warrants further investigation into factors that favorably or adversely affect adoption of ICS by end-users and their perspectives.

### 4.3. Strengths and Limitations

This review has focused on determining the most impactful ICS interventions that will be relevant for the poorest communities globally. Since we restricted the review to studies published from 2012 onwards, this is the first review that looks at the latest evidence on impact of ICS on HAP and BP. Our search strategy was comprehensive in that it included all the major databases for research on air quality as well as health, in addition to a grey literature search. The risk of bias was assessed for all studies and all except one were found to be methodologically strong. To minimize heterogeneity, we restricted the studies to low and middle-income adult populations and follow-up durations not less than 28 days. To control effect modification by pregnancy on BP, we restricted the population of interest to non-pregnant women.

Previous reviews on ICS and BP (Onakomaiya et al, 2019) have included both pregnant and non-pregnant women, this is the first meta-analysis to report impact of ICS interventions on BP solely in non-pregnant women. Despite these restriction criteria and using a random effects model, there was considerable statistical heterogeneity across a variety of sensitivity analyses. Most of the studies, except two were non-randomized studies. Some studies that met the inclusion criteria could not be included in the meta-analysis due to lack of availability of the data in a format that would enable synthesis. This resulted in fewer estimates being used, especially for blood pressure. Lastly, since we restricted the search to literature published in English language, it is possible that other relevant studies published in Spanish, Mandarin or other languages were missed.

## 5. Conclusions and Recommendations

This review shows the greatest reduction of fine particulate matter 2.5 and CO indoor concentration, and blood pressure in the adult population with use of locally made standard combustion cookstoves with chimneys compared to non-chimney cookstoves. This highlights the benefits of designing cookstoves technologies with structural fixes such as chimneys to reduce the household concentration. Also, household energy programs need to further target local artisans to develop ICS that are not only affordable but that meets the local needs of the households. We recommend further research on user’s perspectives on impact of ICS with chimneys as well as qualitative factors that aid or prevent uptake and exclusive use of ICS, for a more comprehensive insight on impact of ICS.

## Data Availability

Data is available upon request.

## Acknowledgements

This review is part of The Smokeless Village Project funded by the Irish Research Council, project number: COALESCE/2020/13. Financial assistance was not sought for this review from the project funding. We are thankful for the institutional support provided by the Royal College of Surgeons in Ireland –Bahrain and Dublin campuses for carrying out this research.

## References

1. Aung TW, Baumgartner J, Jain G, Sethuraman K, Reynolds C, Marshall JD, Brauer M. Effect on blood pressure and eye health symptoms in a climate-financed randomized cookstove intervention study in rural India. Environmental research. 2018 Oct 1;166:658-67.

2. Aung TW, Jain G, Sethuraman K, Baumgartner J, Reynolds C, Grieshop AP, Marshall JD, Brauer M. Health and climate-relevant pollutant concentrations from a carbon-finance approved cookstove intervention in rural India. Environmental science & technology. 2016 Jul 5;50(13):7228–38.

3. Balakrishnan K, Sambandam S, Ghosh S, Mukhopadhyay K, Vaswani M, Arora NK, Jack D, Pillariseti A, Bates MN, Smith KR. Household air pollution exposures of pregnant women receiving advanced combustion cookstoves in India: Implications for intervention. Annals of global health. 2015 May 1;81(3):375–85.

4. Bonjour S, Adair-Rohani H, Wolf J, Bruce NG, Mehta S, Prüss-Ustün A, Lahiff M, Rehfuess EA, Mishra V, Smith KR. Solid fuel use for household cooking: country and regional estimates for 1980– 2010. Environmental health perspectives. 2013 Jul;121(7):784–90.

5. Bruce N, Dherani M, Liu R, Hosgood HD, Sapkota A, Smith KR, et al. Does household use of biomass fuel cause lung cancer? A systematic review and evaluation of the evidence for the GBD 2010 study. Thorax. 2015 May;70(5):433–41. pmid:25758120

6. Clark ML, Bachand AM, Heiderscheidt JM, Yoder SA, Luna B, Volckens J, Koehler KA, Conway S, Reynolds SJ, Peel JL. Impact of a cleaner-burning cookstove intervention on blood pressure in Nicaraguan women. Indoor Air. 2013 Apr;23(2):105–14.

7. Clark SN, Schmidt AM, Carter EM, Schauer JJ, Yang X, Ezzati M, Daskalopoulou SS, Baumgartner J. Longitudinal evaluation of a household energy package on blood pressure, central hemodynamics, and arterial stiffness in China. Environmental research. 2019 Oct 1;177:108592.

8. DerSimonian R, Laird N. Meta-analysis in clinical trials. Controlled clinical trials. 1986 Sep 1;7(3):177–88.

9. Global Alliance for Clean Cookstoves. Handbook for Biomass Cookstove Research, Design, and Development, 2017. Available at: https://www.cleancookingalliance.org/binary-data/RESOURCE/file/000/000/517-1.pdf

10. Goodwin NJ, O’Farrell SE, Jagoe K, Rouse J, Roma E, Biran A, Finkelstein EA. Use of behavior change techniques in clean cooking interventions: a review of the evidence and scorecard of effectiveness. Journal of health communication. 2015 Mar 31;20(sup1):43–54.

11. Hankey S, Sullivan K, Kinnick A, Koskey A, Grande K, Davidson JH, Marshall JD. Using objective measures of stove use and indoor air quality to evaluate a cookstove intervention in rural Uganda. Energy for Sustainable Development. 2015 Apr 1;25:67–74.

12. Higgins JPT, Thompson SG. Quantifying heterogeneity in a meta-analysis. Statistics in Medicine 2002; 21:1539–1558.

13. International Energy Agency (IEA), 2018. World Energy Outlook 2018, IEA, Paris. Available at https://www.iea.org/reports/world-energy-outlook-2018. Accessed June 15, 2020

14. Islam MM, Wathore R, Zerriffi H, Marshall JD, Bailis R, Grieshop AP. In-use emissions from biomass and LPG stoves measured during a large, multi-year cookstove intervention study in rural India. Science of The Total Environment. 2020 Nov 23:143698.

15. Kshirsagar MP, Kalamkar VR. A comprehensive review on biomass cookstoves and a systematic approach for modern cookstove design. Renewable and Sustainable Energy Reviews. 2014 Feb 1;30:580–603.

16. Lea-Langton AR, Spracklen DV, Arnold SR, Conibear LA, Chan J, Mitchell EJ, Jones JM, Williams A. PAH emissions from an African cookstove. Journal of the Energy Institute. 2019 Jun 1;92(3):587–93.

17. Lee KK, Miller MR, Shah AS. Air pollution and stroke. Journal of stroke. 2018 Jan;20(1):2.

18. Milanzi EB, Gehring U. Detrimental effects of air pollution on adult lung function. European Respiratory Journal 2019 54: 1901122. DOI: 10.1183/13993003.01122-2019.

19. Moher D, Liberati A, Tetzlaff J, Altman DG, Prisma Group. Preferred reporting items for systematic reviews and meta-analyses: the PRISMA statement. PLoS med. 2009 Jul 21;6(7):e1000097.

20. Onakomaiya D, Gyamfi J, Iwelunmor J, Opeyemi J, Oluwasanmi M, Obiezu-Umeh C, Dalton M, Nwaozuru U, Ojo T, Vieira D, Ogedegbe G. Implementation of clean cookstove interventions and its effects on blood pressure in low-income and middle-income countries: systematic review. BMJ open. 2019 May 1;9(5): e026517.

21. Pope, D.P., Irving, G., Bruce, N.G., Rehfuess, E.R., 2013. Methodological Quality Assessment for Intervention and Observational Studies using standardised instruments: Liverpool Quality Assessment Tools (LQATs).

22. Puzzolo E, Pope D, Stanistreet D, Rehfuess EA, Bruce NG. Clean fuels for resource-poor settings: a systematic review of barriers and enablers to adoption and sustained use. Environmental research. 2016 Apr 1;146:218-34.

23. Rebelo F, Farias DR, Mendes RH, Schlüssel MM, Kac G. Blood pressure variation throughout pregnancy according to early gestational BMI: a Brazilian cohort. Arquivos brasileiros de cardiologia. 2015 Apr;104(4):284–91.

24. Rehfuess EA, Puzzolo E, Stanistreet D, Pope D, Bruce NG. Enablers and barriers to large-scale uptake of improved solid fuel stoves: a systematic review. Environmental health perspectives. 2014 Feb;122(2):120–30.

25. Sambandam S, Balakrishnan K, Ghosh S, Sadasivam A, Madhav S, Ramasamy R, Samanta M, Mukhopadhyay K, Rehman H, Ramanathan V. Can currently available advanced combustion biomass cook-stoves provide health relevant exposure reductions? Results from initial assessment of select commercial models in India. EcoHealth. 2015 Mar 1;12(1):25–41.

26. Shah AS, Lee KK, McAllister DA, Hunter A, Nair H, Whiteley W, Langrish JP, Newby DE, Mills NL. Short term exposure to air pollution and stroke: systematic review and meta-analysis. BMJ. 2015 Mar 24;350:h1295.

27. Siddharthan T, Grigsby MR, Goodman D, Chowdhury M, Rubinstein A, Irazola V, Gutierrez L, Miranda JJ, Bernabe-Ortiz A, Alam D, Kirenga B. Association between household air pollution exposure and chronic obstructive pulmonary disease outcomes in 13 low-and middle-income country settings. American journal of respiratory and critical care medicine. 2018 Mar 1;197(5):611–20.

28. Singh A, Tuladhar B, Bajracharya K, Pillarisetti A. Assessment of effectiveness of improved cook stoves in reducing indoor air pollution and improving health in Nepal. Energy for sustainable development. 2012 Dec 1;16(4):406–14.

29. Smith K, Mehta S, Maeusezahl-Feuz M. Indoor air pollution from household use of solid fuels. Comp Quantif Heal risks. 2004;1435–93.

30. Stanistreet D, Puzzolo E, Bruce N, Pope D, Rehfuess E. Factors influencing household uptake of improved solid fuel stoves in low-and middle-income countries: A qualitative systematic review. International journal of environmental research and public health. 2014 Aug;11(8):8228–50.

31. United States Agency for International Development (USAID). Technological innovation in cookstoves and fuels. 2017. Available at: https://www.usaid.gov/energy/cookstoves/technological-innovation.

32. Wan X, Wang W, Liu J, Tong T. Estimating the sample mean and standard deviation from the sample size, median, range and/or interquartile range. BMC medical research methodology. 2014 Dec 1;14(1):135.

33. World Health Organization. WHO guidelines for indoor air quality. Household fuel combustion. Geneva, Switzerland 2014. Available at https://www.who.int/airpollution/publications/household-fuel-combustion/en/

34. World Health Organization. Global estimates of burden of disease caused by the environment and occupational risks 2018. Available at: https://www.who.int/news-room/fact-sheets/detail/household-air-pollution-and-health.

35. World Health Organization. Air Pollution, Ambient Air Pollution 2020a. Available at: https://www.who.int/airpollution/ambient/pollutants/en/.

36. Yuan S, Xu W, Liu Z. A study on the model for heating influence on PM2. 5 emission in Beijing China. Procedia Engineering. 2015 Jan 1;121:612–20.

