## Supplementary Material for "Do improved biomass cookstove interventions improve indoor air quality and blood pressure? A systematic review and meta-analysis"

### S1 Keywords for Study Search

| | | |
|---|---|---|
| | **SCOPUS**<br><br>Elsevier.com | |
| 1 | ( TITLE-ABS-KEY (fuel W/1 stove* OR fuelstove* cookstove* OR cook W/1 stove* OR cookingstove* OR cooking W/1 stove* OR cooker OR cookers ) ) ) | 663 |
| 2 | ( TITLE-ABS-KEY ( air W/1 pollution OR air W/1 pollutant* OR air W/1 quality ) ) | 90799 |
| 3 | ( TITLE-ABS-KEY ( household OR households OR family OR families OR home OR homes OR homestead* OR domestic OR residential ) ) | 2958925 |
| 4 | #1 AND #2 AND #3 | 71 |
| | **Global Health Database on OVID** | |
| 1 | (fuel adj1 stove*) OR fuelstove* OR cookstove* OR (cook adj1 stove*) OR cookingstove* OR (cooking adj1 stove*) OR cooker OR cookers | 613 |
| 2 | exp air pollution/ OR exp indoor air pollution/ OR (air adj1 pollution) OR (air adj1 pollutant*) OR (air adj1 quality) | 27509 |
| 3 | household OR households OR family OR families OR home OR homes OR homestead* OR domestic OR residential | 286386 |
| 4 | #1 AND #2 AND #3 | 206 |
| 5 | LIMIT #5 2000-2020 | 199 |
| | **PubMed**<br><br>at https://pubmed.ncbi.nlm.nih.gov/ | |
| 1 | stove OR stoves OR "fuel stove" OR fuelstove OR cookstove* OR "cook stove" OR cooking OR cookingstove* OR "cooking stove" OR cooker OR cookers | 31,130 |
| 2 | "Air Pollutants"[Mesh] OR "Air Pollution"[Mesh] OR "Air Quality"[Mesh] OR "Air Pollution, Indoor"[Mesh] OR "air pollution" OR "air pollutant" OR "air quality" | 116,732 |
| 3 | (household OR households OR family OR families OR home OR homes OR homestead* OR domestic OR residential)[Text Word] | 1,870,42<br>3 |
| 4 | #1 AND #2 AND #3 | 1,560 |
| 5 | Limit #4 to Humans and 2010 to 2020 | 729 |
| | **Web of Science**<br>**Science, Social Science, Conference Proceedings, Book Index 1945-2020** | |
| 1 | TS=( fuel NEAR/1 stove* OR fuelstove* OR cookstove* OR cook NEAR/1 stove* OR cookingstove* OR cooking NEAR/1 stove* OR cooker OR cookers) | 1569 |
| 2 | TS=(air NEAR/1 pollution OR air NEAR/1 pollutant* OR air NEAR/1 quality) | 117,410 |
| 3 | TS=(household OR households OR family OR families OR home OR homes OR homestead* OR domestic OR residential) | 2,041,93<br>7 |
| 4 | #1 AND #2 AND #3 | 612 |
| 5 | Limit #4 to 2010 to 2020 | 526 |
| | **EMBASE**<br>Elsevier.com | |

| | | |
|---|---|---|
| 1 | ( fuel NEXT/1 stove* OR fuelstove* OR cookstove* OR cook NEXT/1 stove* OR cookingstove* OR cooking NEXT/1 stove* OR cooker OR cookers):ti,ab,kw | 1216 |
| 2 | (air NEXT/1 pollution OR air NEXT/1 pollutant* OR air NEXT/1 quality) | 56118 |
| 3 | (household OR households OR family OR families OR home OR homes OR homestead* OR domestic OR residential):ti,ab,kw | 1651217 |
| 4 | #1 AND #2 AND #3 | 337 |
| 5 | Limit #4 to 2010 to 2020 and exclude Medline records | 102 |
| | **Cochrane Library and Register of Clinical Trials**<br>Wiley.com | |
| 1 | stove OR stoves OR "fuel stove" OR fuelstove OR fuelstoves OR cookstove* OR "cook stove" OR cooking OR cookingstove* OR "cooking stove" OR cooker OR cookers | 1430 |
| 2 | air NEAR/1 pollution OR air NEAR/1 pollutant* OR air NEAR/1 quality | 1331 |
| 3 | household OR households OR family OR families OR home OR homes OR homestead* OR domestic OR residential | 87878 |
| 4 | #1 AND #2 AND #3 | 9 reviews & 145 trials |
| 5 | Limit #4 to 2010 - 2020 | 9 reviews & 136 trials |
| 6 | Selected 9 reviews and not trails of clinical interventions for related disease | **9** |
| | **LILACS Informação em Saúde da América Latina e Caribe**<br><br>https://pesquisa.bvsalud.org/portal/ Advanced search | |
| 1 | (tw:( stove OR stoves OR fuelstove OR fuelstoves OR cookstove OR cooking OR cookingstove ... | 1450 |
| 2 | Filtered to exclude Medline and limited to 2010-2020 | 12 |
| | **AFRICA BIB**<br>https://www.africabib.org/ | |
| | (Stove OR stoves OR fuelstove OR fuelstoves OR cookstoves OR cooking OR cookingstove OR cooker OR cookers) AND (pollution) | 0 |
| | **BASE Biefeld Research search engine**<br>https://www.base-search.net/ | |
| | (Stove OR stoves OR fuelstove OR fuelstoves OR cookstoves OR cooking OR cookingstove OR cooker OR cookers) AND (air) AND (pollution) LIMITED to most recent 200<br><br>319 hits in 164,715,446 documents when limited to books, reports, reviews, conference papers, theses | 200 |
| | **CAMPBELL COLLABORATION**<br>https://campbellcollaboration.org/ | |
| 1 | (stove OR stoves OR "fuel stove" OR fuelstove OR fuulstoves OR cookstove* OR "cook stove" OR cooking OR cookingstove* OR "cooking stove" OR cooker OR cookers) | 0 |
| 2 | (air pollution OR air pollutants OR air quality) | 0 |

### S2 Data Extraction Template

| 1 | # | Outcome | Unit | Authors, Title, Yr | Design | Stove type | Stove brand | Duration (days) | Location | Intervention pre 'N' | Intvn pre Mean | Intvn pre SD | Intvn post 'N' | Intvn post mean | Intvn post SD |
|---|---|---|---|---|---|---|---|---|---|---|---|---|---|---|---|
| 2 | | | | | | | | | | | | | | | |
| 3 | | | | | | | | | | | | | | | |
| 4 | | | | | | | | | | | | | | | |
| 5 | | | | | | | | | | | | | | | |
| 6 | | | | | | | | | | | | | | | |
| 7 | | | | | | | | | | | | | | | |
| 8 | | | | | | | | | | | | | | | |
| 9 | | | | | | | | | | | | | | | |

**S3 Example of Risk of Bias assessment using Liverpool Quality Assessment Tool (LQAT)**

| Liverpool University Quality Assessment Tool (LQAT) | | | | |
|---|---|---|---|---|
| **Study ID (Author, year and date of extraction):** **Alexander et al., 2016. Extraction: 9-2020**<br>**Study design:** Randomized Controlled Trial<br>**Brief – study methods:** Aim: evaluate the effect of a clean cookstove intervention on blood pressure during pregnancy. Intervention implemented in Nigeria. Pregnant women cooking with kerosene or firewood were randomly assigned to an ethanol or control arm. Blood pressure measurements were taken during six month antenatal visits. Intervention versus controls were compared in the primary analysis. In subgroup analyses, baseline kerosene users assigned to the intervention were compared with kerosene control subjects and baseline firewood users assigned to the intervention were compared with firewood control subjects. | | | | |
| | **WEAK** | **MODERATE** | **STRONG** | **REASON and IMPLICATION** |
| **SELECTION PROCEDURES (population/sample size, sampling method)** | | | X | Pregnant women screened for eligibility at four primary healthcare centres. 324 women randomly assigned to IG or CG. Convenience sample, sample size determination not discussed. |
| **BASELINE ASSESSMENT (baseline fuel/stove intervention details and how distributed)** | | | X | Intervention: women given ICS + initial supply of fuel during a home visit. Comprehensive training regarding smoke exposure + proper use of stove provided. Staff observed participants use ICS for the first time. CG received education on dangers of smoke exposure. |
| **OUTCOME ASSESSMENT (assessment of scaling up/adoption/use)** | | | X | Assessment only measured effects on BP. No measurement/assessment of scaling up/adoption/use. |
| **ANALYSIS/CONFOUNDING (how data analysed/presented)** | | | X | Analyses between ICS and TCS compared, as well as secondary analyses stratified based on pre-intervention stove type. |
| **IMPACT (applicability/impact of findings to review)** | | | X | Improved DBP, but not SBP. Unclear about participant's opinion on stove use. |
| GLOBAL RATING SCORE (SEE score key BELOW) | | | | **Strong** |

**GLOBAL RATING FOR THIS PAPER**

STRONG ____(no WEAK ratings)
MODERATE ________(one WEAK rating)
WEAK (two or more WEAK ratings)
